# Capture of Group A Streptococcus by Open-Microfluidic CandyCollect Device in Pediatric Patients

**DOI:** 10.1101/2023.12.14.23299923

**Authors:** Wan-chen Tu, Ingrid H. Robertson, Andrea Blom, Elena Alfaro, Victoria A. M. Shinkawa, Daniel B. Hatchett, Juan C. Sanchez, Anika M. McManamen, Xiaojing Su, Erwin Berthier, Sanitta Thongpang, Ellen R. Wald, Gregory P. DeMuri, Ashleigh B. Theberge

**Affiliations:** Department of Chemistry, University of Washington, Seattle, WA, USA; Department of Pediatrics, University of Wisconsin School of Medicine and Public Health, Madison, WI, USA; Department of Biomedical Engineering, Faculty of Engineering, Mahidol University, Nakorn Pathom, Thailand; Department of Urology, School of Medicine, University of Washington, Seattle, WA, USA

**Keywords:** Pediatric diagnostics, Group A Streptococcus, patient centric sample collection, point-of-care, open microfluidics

## Abstract

**State the purpose:** Obtaining high-quality samples to diagnose streptococcal pharyngitis in pediatric patients is challenging due to discomfort associated with traditional pharyngeal swabs. This may cause reluctance to go to the clinic, inaccurate diagnosis, or inappropriate treatment for children with sore throat. Here, we determined the efficacy of CandyCollect, a lollipop-inspired open-microfluidic pathogen collection device, to capture Group A Streptococcus (GAS) and compare user preference for CandyCollect, conventional pharyngeal swabs, or mouth swabs in children with pharyngitis and their caregivers.

**Results:** All child participants (30/30) were positive for GAS by qPCR on both the mouth swab and CandyCollect. Caregivers ranked CandyCollect as a good sampling method overall (27/30), and all caregivers (30/30) would recommend CandyCollect for children 5 years and older. Twenty-three of 30 children “really like” the taste and 24/30 would prefer to use CandyCollect if a future test were needed. All caregivers (30/30) and most children (28/30) would be willing to use CandyCollect at home.

**Conclusion:** All participants tested positive for GAS on all three collection methods (pharyngeal swab, mouth swab, and CandyCollect). While both caregivers and children like CandyCollect, some caregivers would prefer a shorter collection time. Future work includes additional studies with larger cohorts presenting with pharyngitis of unknown etiology and shortening collection time while maintaining the attractive form of the device.

**Translational Impact Statement:** Obtaining oral samples for the diagnosis of streptococcal pharyngitis is of great importance for children. To address the challenges associated with traditional pharyngeal swab sampling, we developed the CandyCollect device, a lollipop-inspired open mesofluidic saliva sampling system. In this study, saliva samples were collected from children, aged 5-14 years, with CandyCollect and mouth swabs and analyzed via qPCR. The results show CandyCollect is the child preferred collection tool and had 100% concordance with the results from traditional diagnosis methods as part of their clinical care.

## Introduction

Group A Streptococcus (GAS) accounts for over 600 million cases of bacterial pharyngitis worldwide each year,^1^ affecting 15% of school-aged children in developed countries. In developing nations the burden is markedly greater, with rates 5-10 times higher.^1^ Though symptoms of GAS pharyngitis almost always resolve after 3-4 days, antibiotics are recommended to shorten the course of the infection, lessen spread, and prevent acute suppurative complications.^1^ Accurate detection of GAS is imperative; untreated and recurring GAS infections can lead to severe outcomes, such as acute rheumatic fever, which causes carditis in 30-45% of cases and is the most common cause of acquired heart disease in children worldwide.^1^

A standard method for diagnosing GAS pharyngitis is with a rapid antigen detection test (RADT), which requires a posterior pharyngeal swab sample and provides a positive or negative result within minutes.^2^ If the RADT is negative, a throat culture is performed on a posterior pharyngeal swab in order to detect false negatives, and laboratory analysis takes more time.^2,3^ Another option is a molecular test (e.g., quantitative polymerase chain reaction (qPCR)), again requiring a posterior pharyngeal swab.^4^ Pharyngeal swabs are uncomfortable, often acting as a deterrent for children and adults; however, it is important to confidently diagnose bacterial pharyngitis not only for the long-term health of the patient but also to avoid over-prescribing antibiotics.^1^ DeMuri, et al. established that mouth swabs (sucked on like a lollipop) provide an adequate sample for the detection of GAS when sensitive methods such as PCR are used.^5^

With developments in PCR and RADTs using saliva instead of posterior pharyngeal swabs, more options are now available for salivary detection, such as spit tubes (OMNIgene™), passive drooling, mouth swabs (Eswab™), and cotton rolls (Salivette®).^6–8^ The CandyCollect, a lollipop-inspired open-fluidic sampling device, was first introduced in our 2021 in-lab study that showed its ability to capture and accumulate *Streptococcus pyogenes* for analysis.^9^ In 2022, we tested the device with a remote, at-home human subjects study in which the CandyCollect devices effectively captured commensal bacteria (*Streptococcus mutans* and *Staphylococcus aureus*) from healthy adults.^10^ The CandyCollect devices were shipped to the lab in dry tubes under ambient conditions to be quantified using qPCR from the elution fluid.^10^ Ours is not the only lollipop-inspired device; there are at least three other collection or testing devices with a similar lollipop form already on the market, although these alternative devices do not look like a traditional candy lollipop and also do not provide flavor. The Self-LolliSponge™ by Copan has the user keep the sponge stick in their mouth for a “few minutes” with a lemon scented cap to improve the experience.^11^ The V-Chek COVID-19 Saliva Antigen Test provides a “‘Lollipop’ Saliva Swab” that the user places in their mouth for 90 seconds and then attaches to a lateral flow assay cassette.^12^ The Whistling COVID-19 Saliva Antigen Rapid Test combines an absorbent material attached to a handle that the user places in their mouth with an immunochromatographic assay.^12^

Obtaining high-quality samples to diagnose GAS pharyngitis in pediatric patients is important for treatment; however, the current collection tools are not child friendly. Here we aimed to demonstrate the feasibility of the CandyCollect device by collecting salivary samples from 30 pediatric patients aged 5-17 years with known GAS pharyngitis. Further we aimed to compare user preference for CandyCollect, conventional pharyngeal swabs, or mouth swabs among children with pharyngitis and their caregivers.

## Method

### Mouth swabs and CandyCollect devices

#### Swabs and media

The pharyngeal swab used in the clinical visit was a standard of care double swab, BBL™ CultureSwab™ Transport Systems by Copan (FisherScientific, Cat# B4320109). The white-capped Eswab™ with nylon flocked swabs and Liquid Amies media collection kits by Copan (FisherScientific, Cat# R723480) were used as mouth swabs.

#### CandyCollect device stick fabrication

The CandyCollect device is milled from a sheet of 4 mm polystyrene (Goodfellow, Cat# 700-272-86) using the DATRON Neo computerized numerical control (CNC) mill as previously described.^9^ The Fusion file and an engineering drawing of the CandyCollect device are included in the Supplemental Material (Supplemental Figure 1). After sanding rough edges, devices were sonicated in isopropanol (IPA) (FisherScientific, A451-4) and 70% v/v ethanol (FisherScientific, Decon™ Labs, 07-678-004).

#### Plasma treatment of CandyCollect devices

A Zepto LC PC Plasma Treater (Diener Electronic GmbH, Ebhausen, Germany), using oxygen gas, increased hydrophilicity of the device surface. The process includes decreasing the chamber pressure to 0.20-0.25 mbar and adding oxygen gas while 70 W voltage is applied for 5 minutes.^9,10^

#### Preparation of CandyCollect devices for human subjects study

CandyCollect devices were prepared as previously described with the exception of storage containers and candy mass.^9,10^ Half of the patients (1-15) were sampled with CandyCollects made with 1.2-1.34 g isomalt candy, while the second half (16-30) were sampled with 0.45-0.57 g; procedures are duplicated here for convenience. In accordance with the kitchen hygiene guidelines of the Washington State Cottage Food Operations Law (RCW 69.22.040(2b-f(ii-iv))), candy was applied to CandyCollect sticks by lab members who are trained in food safety, have a Food Worker Card (WA State), and wore gloves and a mask during food preparation.

Candy was prepared by gradually adding isomalt to water while heating to a target temperature of 171°C, then adding food coloring. Next, the isomalt mixture was cooled in a room temperature water bath while strawberry candy flavoring was added. Plasma treated CandyCollect sticks were cleaned using hot water and dish soap. After the sticks were dry, small amounts of isomalt candy were re-melted and added to silicone molds with the CandyCollect sticks. Once the candy was cooled, the mass was recorded, and the device was heat-sealed in a polypropylene bag. Devices were stored in Ziploc® storage bags with a food safe desiccant (Amazon, Cat# B00DYKTS9C) until being sent to UW-Madison for the clinical study.

### Clinical Study Procedures

#### Subjects and Specimens

Caregiver-child dyads were recruited from an ambulatory care clinic that serves children in Madison, Wisconsin. Children were diagnosed with GAS pharyngitis using a traditional pharyngeal swab via RADT. The study was approved by the UW-Madison Health Sciences Institutional Review Board (#2021-1427). Study data were collected and managed using REDCap (Research Electronic Data Capture) hosted at the University of Wisconsin-Madison, School of Medicine and Public Health.^13,14^

A parent or legal guardian (aka “caregiver”) provided verbal consent for their own participation in the study and caregiver permission for their child, and all children provided verbal assent. Eligible children needed to be able to suck on swabs and the CandyCollect devices and could not have a sensitivity to sugar-free products; additionally, a parent or legal guardian needed to be present and enroll in the study along with the child.

Immediately after the clinic visit, in which the traditional pharyngeal swab was obtained, a research nurse guided participants through collecting four oral samples: two commercially available mouth swabs (Eswab™) for 10 seconds each and two CandyCollect devices until the candy was fully dissolved (Figure 1). The samples were stored at −80°C and shipped on dry ice prior to qPCR analysis. Race and ethnicity information was collected based on the requirements of NIH. Race and ethnicity information was taken from the electronic health record, where it had previously been entered based on self-report by the parent/legal guardian.

**Figure 1.**
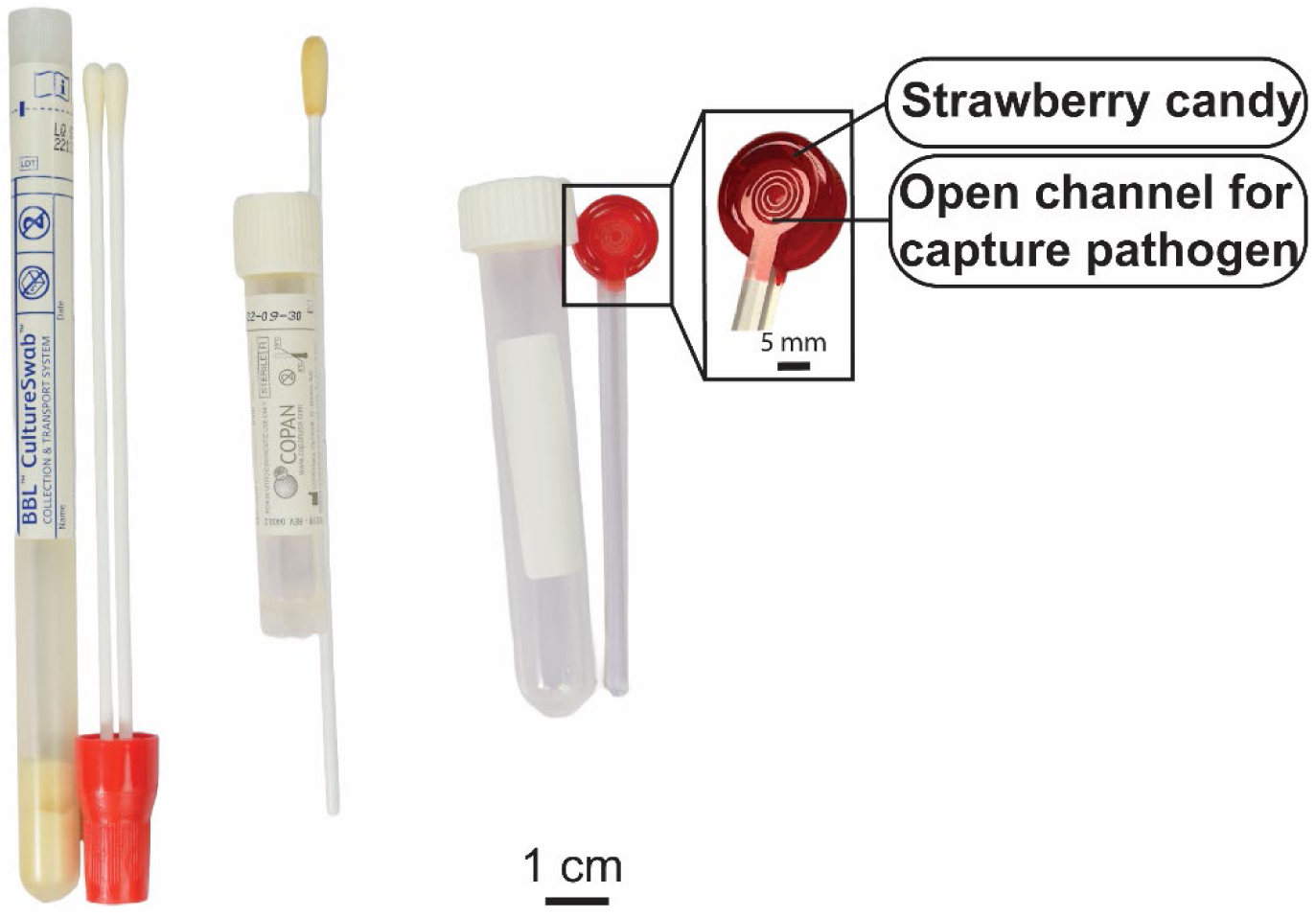
GAS sampling tools used in this study: pharyngeal (throat) swab, ESwab™ (mouth swab), and the CandyCollect device (from left to right). The components of the CandyCollect device include: 1) strawberry candy to facilitate pleasant user experience of the CandyCollect device and 2) open channels which capture bacterial pathogens.

#### Testing Preference Survey

After sample collection was complete, the research nurse gave a paper-based survey to both the child and their caregiver. All survey questions are included in supplementary materials, Supplemental Table 1-2. The research nurse provided instructions about the Wong-Baker FACES® Pain Rating Scale to all child participants, explaining that each face represents a person who has no pain (hurt), or some, or a lot of pain (Supplemental Figure 2).^15^ The nurse asked the participants to choose the face that best describes the pain they experienced during each type of testing. The research nurse assisted children with completing the form if they needed help reading or understanding. The research nurse gave caregivers privacy in completing their surveys, to encourage honest responses, only answering questions as needed. Participants were also asked to provide suggestions and free responses about the main reasons for their preferences. All free response questions are included in Supplemental Table 3.

### Specimen Processing and Laboratory Analysis

The detailed specimen processing and laboratory analysis procedures were previously described.^10^ In brief, the mouth swab and CandyCollect sample collection was directed by a research nurse. Samples were stored in −20 °C and then transferred to storage at −80 °C within 12 days. Samples were shipped to the University of Washington on dry ice and then stored at −80 °C before processing. All laboratory procedures were performed in accordance with Biosafety Level-2 laboratory practices and the University of Washington Site-Specific Bloodborne Pathogen Exposure Control Plan. CandyCollect devices and mouth swabs were processed as previously described;^10^ additional details are in the Supplemental Material. Briefly, the material on the CandyCollect device was eluted by phosphate-buffered saline (PBS) (Gibco™, Cat# 10010023) with 5% Proteinase K (Thermo Scientific™, Cat# EO0491); for mouth swab samples, *S. pyogenes* DNA was isolated using MagMAX™ Total Nucleic Acid Isolation Kit (ThermoFisher Scientific, Cat# AM1840) according to the protocol supplied by the manufacturer. After elution and lysis, the samples were also enriched and purified using MagMAX™ Total Nucleic Acid Isolation Kit following the protocol supplied by the manufacturer. ^10^

qPCR was performed for the detection of *S. pyogenes* genomic DNA as previously described.^9, 10^ The results were expressed as Cycle threshold (Ct, number of cycles until the sample was detected above the threshold). Any Ct value below or equal to Ct values for the lowest concentration of DNA standards (5 fg) on the same plate was considered positive; this cut-off was determined from standard curves performed with genomic DNA isolated from *S. pyogenes*.^9^

Statistics: Statistical analysis was performed using GraphPad Prism 9 software. Paired t-tests were used to evaluate the significance of pairwise comparisons.

### Preparation of positive and negative controls

Preparation of positive controls (via in-lab capture of *S. pyogenes*) and negative controls is described in our prior publications ^9, 10^ and the Supplemental Material. In brief, *S. pyogenes* was prepared from *Streptococcus pyogenes* Rosenbach (American Type Culture Collection, ATCC®, Cat# 700294™), and cultured in Todd-Hewitt Yeast broth.^16^ *S. pyogenes* culture was inoculated one day prior to experiments. For positive controls, 50 µL of *S. pyogenes* culture media were added to the CandyCollect devices, and mouth swabs were soaked in the *S. pyogenes* culture media for 10 seconds. PBS was used as the negative control sample.

## Results

### Demographic Characteristics

In this study, 30 pediatric child-caregiver dyads were enrolled at a clinic in Madison, Wisconsin. Enrolled child participants were aged 5 to 14 years with GAS pharyngitis, which is consistent with the age range in which GAS pharyngitis is most common;^1^ the mean age of participants was 8.8 years (Figure 2 and Supplemental Table 4). More than 90% of child participants were white and not Hispanic. (Supplemental Table 4).

**Figure 2.**
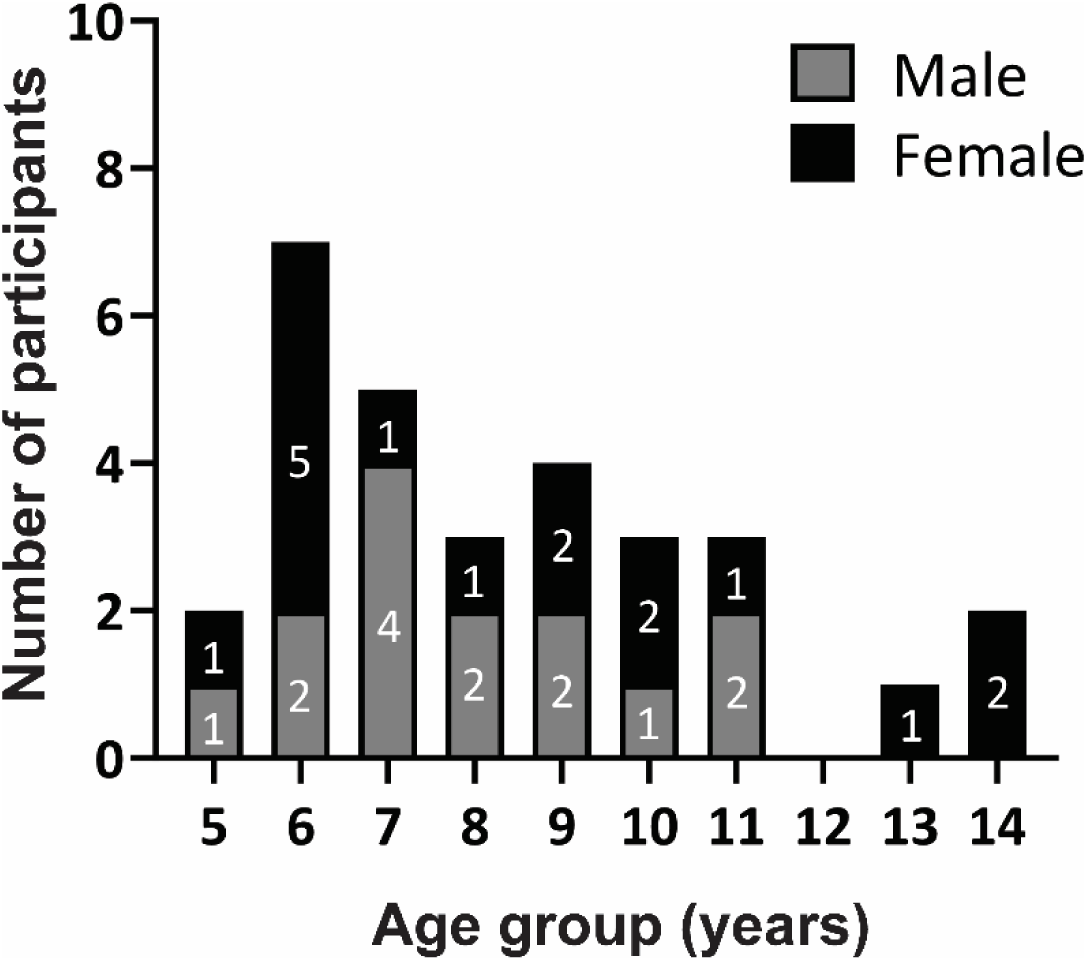
Subject demographics. Thirty participants, aged 5-14 years, were enrolled in this study. Of all the participants, 16 were females and 14 were males.

### qPCR analysis of GAS in CandyCollect devices and mouth swabs

Each sample was analyzed using qPCR and the Ct is shown in Figure 3. The samples collected by CandyCollect devices have an average Ct of 31.28 (range=19.1-38.6) and mouth swabs have an average Ct of 32.08 (range=19.5-38.1); the Ct from the two sampling methods of the 30 participants are not significantly different (p>0.05). Nine samples collected by CandyCollect devices have a higher Ct than the corresponding mouth swabs and the Ct of 13 samples collected by CandyCollect devices is lower than the corresponding mouth swabs. Two different versions of the CandyCollect device (with 0.5 g and 1.3 g candy) were used, with half of the participants using each size. The comparison of the Ct values from two different sizes of candy (15 participants each size), shown in Supplemental Figure 3, are not significantly different (p>0.05). There is also no notable correlation between the CandyCollect device consumption time and the Ct values (Figure 4).

**Figure 3.**
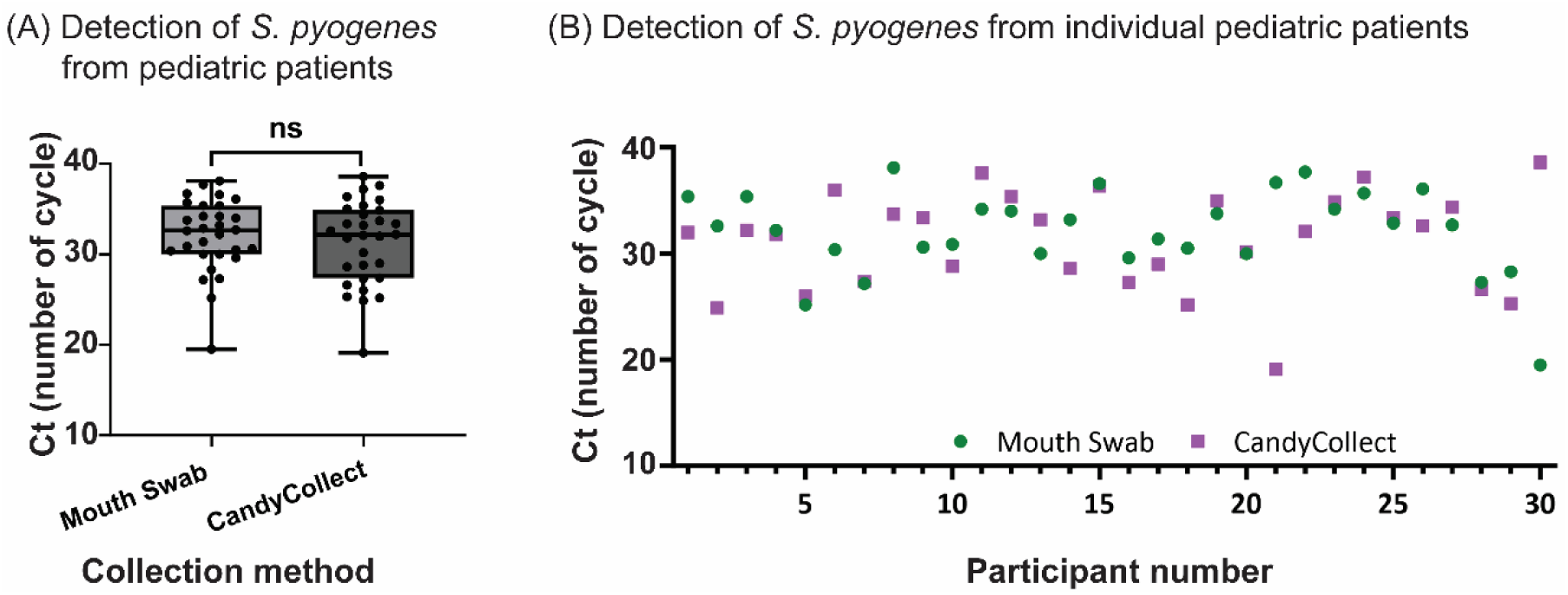
(A) Pooled cycle threshold (Ct) values of samples collected from 30 child participants with GAS pharyngitis with the mouth swabs (left, light grey) and CandyCollect devices ((right, dark grey). Ct from the two sampling methods of all 30 participants are not significantly different (p>0.05). (B) individual Ct values collected from the CandyCollect devices (purple, square) and mouth swabs (green, circle). Data are plotted by the Ct of the CandyCollect devices; the participant numbers were randomly re-numbered. Any Ct value below or equal to Ct values for the lowest concentration of DNA standards (5 fg) on the same plate was considered positive; this cut-off was determined from standard curves prepared with genomic DNA isolated from *S. pyogenes*; all points plotted are positive results.

**Figure 4.**
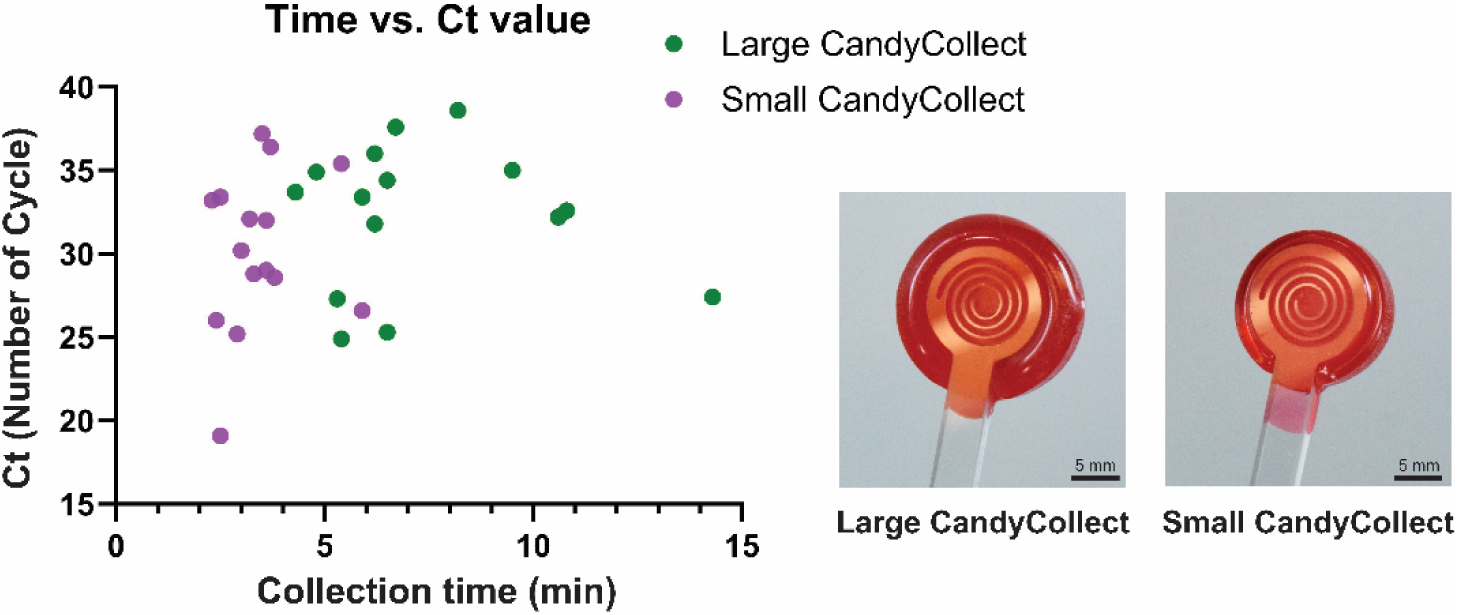
Collection time of CandyCollect devices vs. Ct values of samples collected from 15 participants using the large CandyCollect devices (Candy = 1.3 g, green) and from 15 participants using small CandyCollect devices (Candy = 0.5 g, purple).

### User Feedback of CandyCollect devices and two other commercial collection tools

All 30 children and their caregivers completed feedback surveys. Twenty-nine of 30 children responded that they did not feel any pain or discomfort while using the CandyCollect devices; in comparison, more than half of participants felt pain/discomfort with the traditional throat swabs (Figure 5). Twenty-seven of 30 caregivers ranked the CandyCollect as a good sampling method overall; 29/30 caregivers indicated that using the CandyCollect device was a pleasant experience for their child (Figure 5). The majority of children reported that the CandyCollect device is easy to use (17/30), and they “really like” the taste (n= 23/30) (Figure 6). The majority of caregivers reported that the appearance of the CandyCollect was appealing (n= 26/30) (Figure 6). When asked to consider using CandyCollect, mouth swab, or throat swab if they needed another GAS pharyngitis diagnostic test next week, more than a half of the children (24/30) and caregivers (17/30) preferred to use the CandyCollect device, with the remaining children and caregivers preferring the mouth swab; no participants selected the throat swab (Figure 6). For caregivers of children who consumed the 0.5 g CandyCollect device (consumption time range 2.3-5.9 min; 3.3 min median), 12/15 preferred the CandyCollect device and 3/15 preferred the mouth swab; for caregivers of children who consumed the 1.3 g CandyCollect device (consumption time range 4.3-14.2 min; 6.5 min median), 5/15 preferred the CandyCollect device and 10/15 preferred the mouth swab. Almost all children (28 out of 30) and all caregivers (30 out of 30) stated that they would be willing to use the CandyCollect device at home (Figure 6). All caregivers report they would recommend using the CandyCollect device (Figure 6) for children 5 years and older. All survey questions are included in Supplemental Table 1-2.

**Figure 5.**
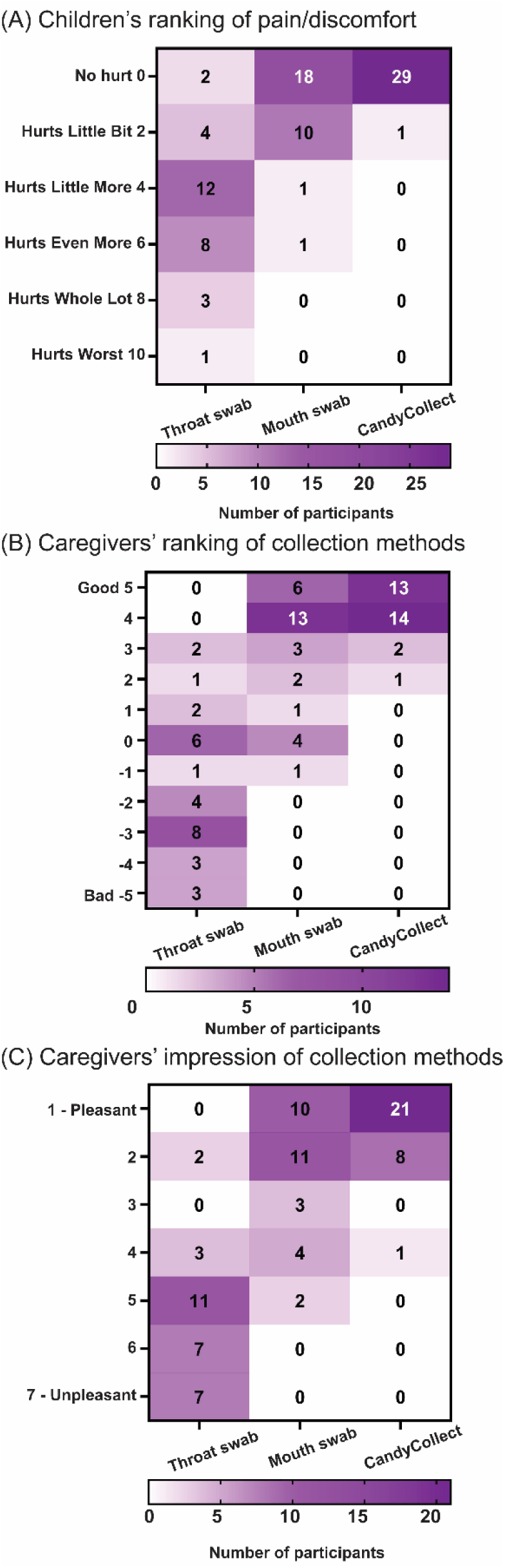
Survey responses to assess the comparison of three collection methods. All survey questions are included in Supplemental Table 1 and 2: (A) Supplemental Table 1 questions 1-3 (B) Supplemental Table 2 questions 2, 4, and 6 (C) Supplemental Table 2 questions 1, 3, and 5.

**Figure 6.**
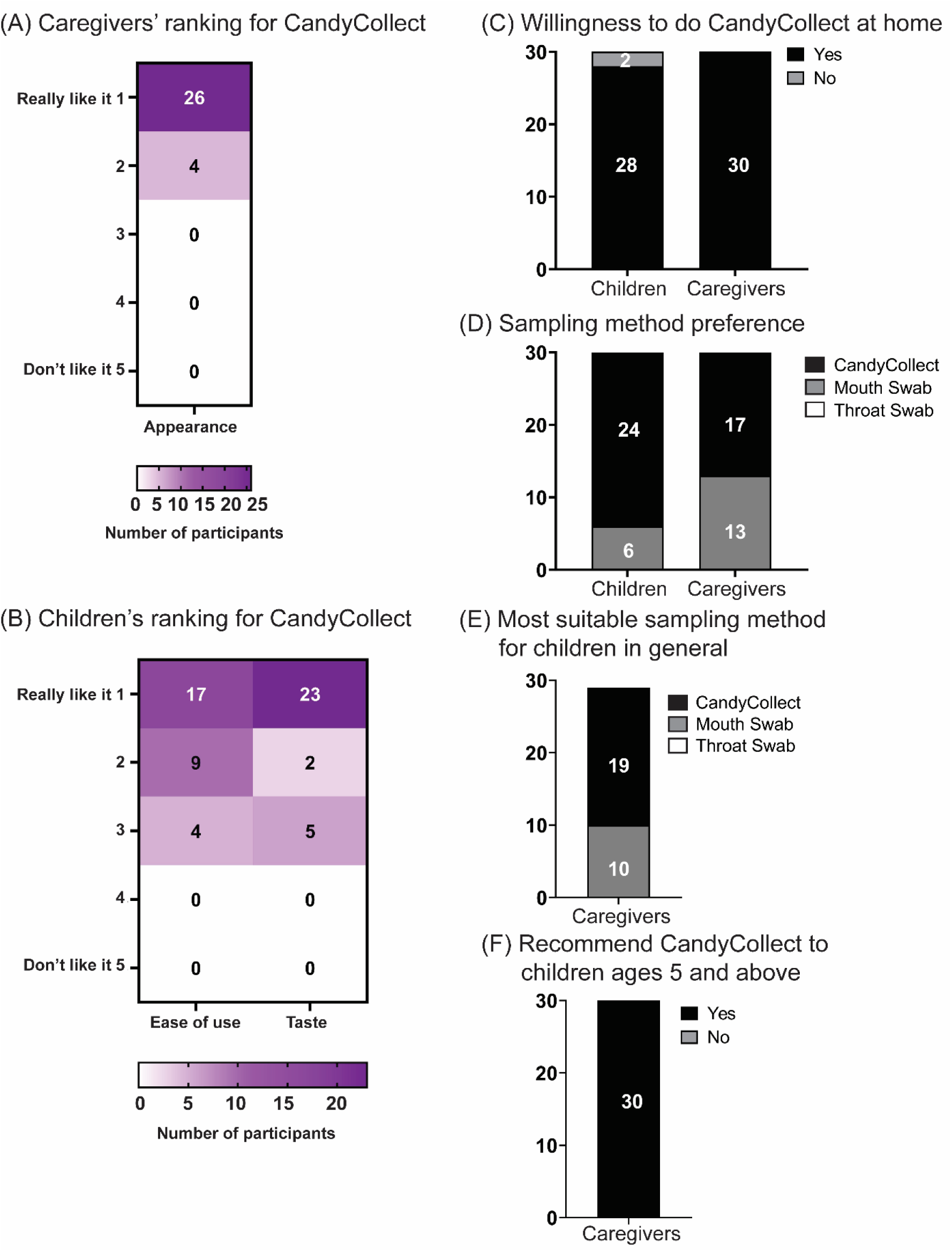
Survey responses to assess the ranking of CandyCollect devices and user preferences. All survey questions are included in Supplemental Table 1 and 2. (A) Supplemental Table 2 question 10 (B) Supplemental Table 1 questions 6 and 7 (C) Supplemental Table 1 question 5 and Supplemental Table 2 question 9 (D) Supplemental Table 1 question 4 and Supplemental Table 2 question 7 (E) Supplemental Table 2 question 8 (F) Supplemental Table 2 question 11.

## Discussion

The ultimate goal of the CandyCollect device is to capture bacterial pathogens for laboratory detection. In our previous in-lab study, we developed the CandyCollect device to capture *S. pyogenes* in saliva and established a qPCR assay to detect *S. pyogenes* eluted from CandyCollect devices.^9^ We also used CandyCollect devices to capture commensal bacteria (*Streptococcus mutans* and *Staphylococcus aureus*) from healthy adults in a home setting as proof-of-concept human subjects research.^10^ The present publication is the first human subjects research that used CandyCollect devices with pediatric patients. The results show that all 30 children diagnosed with GAS pharyngitis using a RADT were positive on CandyCollect devices and mouth swabs. As expected in studies of human subjects, there was some variability in the amount of bacteria (Ct value) across sampling methods and participants, which was also shown in a previous study comparing throat swabs and mouth swabs.^5^

From the user feedback surveys, many caregivers noted the CandyCollect device is less frightening for children than other sampling methods, especially in comparison to throat swabs. Several caregivers noted that the CandyCollect device is easy, fun, and non-invasive. Some caregivers indicated that the CandyCollect device could help children be less afraid of going to the doctor or even be excited to do the test. All survey free responses and suggestions are included in Supplemental Table 3. When asked for suggestions to improve CandyCollect devices, the most common suggestion was to shorten the sampling time. We responded to this suggestion mid-study by using a CandyCollect device that afforded a shorter sampling time for the second half of participants. As noted in the Results, caregivers indicated a preference for either the CandyCollect device or the mouth swab, with no caregivers (and no children) preferring the throat swab. Notably, more caregivers preferred their children to use CandyCollect devices rather than mouth swabs when their child used smaller CandyCollect devices with a faster dissolving time (12/15) in comparison to caregivers of children who used the larger CandyCollect devices with a longer dissolving time (5/15). Slightly more children also preferred using CandyCollect devices when they used faster-dissolving CandyCollect devices (13/15 vs. 11/15). Schuster et al., also reported the same reasoning for sampling preferences: 73% of participants preferred the nasal swab compared to saliva collection by drooling due to the faster sampling time.^17^ We are currently working to modify the CandyCollect device so as to have faster dissolving candy.

There are three major limitations for this study. First, this pilot study was performed with a relatively small sample size; we have ongoing studies with larger sample sizes in adults and children with GAS and other respiratory illnesses. Second, participants were recruited from a single site with relatively homogenous demographics, although we have no reason to suspect that demographics influence capture of pathogens in saliva. Our ongoing work includes remote nationwide studies with enrollment demographics matching national distribution of race/ethnicities. Finally, to maximize the number of positive cases tested with CandyCollect devices in this feasibility study, we only included children who tested positive for GAS on pharyngeal swabs analyzed with RADT during their clinic visits; future studies will include children with pharyngitis of unknown etiology evaluated with standard clinical and research protocols.

### Conclusion

The CandyCollect device, a user-friendly pathogen collection method, is specifically designed to make both in-clinic and at-home sampling more accessible for children. In our inaugural pediatric clinical study, we demonstrated the CandyCollect device successfully captured GAS from 30 children in a clinical setting. Notably, a majority of the children and their caregivers expressed a preference for the CandyCollect device as their chosen method for collecting GAS (compared to traditional throat swabs and mouth swabs). This study represents a significant step towards facilitating collection of samples for the detection of GAS and other respiratory pathogens. Currently, we are modifying the CandyCollect device based on user feedback to optimize its performance and user experience, and we will enroll more participants to achieve larger sample sizes of children with GAS pharyngitis. The present study paves the way for several exciting research directions, including: (1) integrating existing lateral flow immunoassay techniques with the CandyCollect device to enhance the diagnostic capabilities at home and shorten the diagnostic time, and (2) expanding the range of pathogens that can be captured using the CandyCollect device.

## Data Availability

All data produced in the present study are available upon reasonable request to the authors.

## Acknowledgments

This work was supported by National Institutes of Health grants (R21AI166120, R35GM128648 (the latter specifically supported some of the in-lab developments and procedure developments)); the Washington Research Foundation; the Camille and Henry Dreyfus Foundation; and an Alfred P. Sloan Research Fellowship. The content is solely the responsibility of the authors and does not necessarily represent the official views of the National Institutes of Health or other funding sources.

We would like to thank Jason Robertson for his review of the manuscript; Lochlan Hickok and Paul Miller for helping facilitate shipping and logistics; Karen N. Adams advising on work with human subjects study design and regulatory protocols; Alexandra Lindstrom and Bridget L. Johnson advising on study design and execution. Moreover, we thank the participants enrolled in this study.

## Author contributions

W-c. T. designed the sample processing protocol, carried out the sample analysis, performed data analysis, made figures for the manuscript, drafted the initial manuscript, and critically reviewed and revised the manuscript. I. J. fabricated the CandyCollect devices and prepared devices for human subject research, interpreted the data, drafted the initial manuscript, and critically reviewed and revised the manuscript. A. B. advised on study design and execution, enrolled and interacted with the participants, collected the samples and user surveys, accessioned and shipped the samples, interpreted the data, and critically reviewed and revised the manuscript. E. A. advised on study design and execution, advised on all regulatory aspects, drafted the study protocol and survey instruments, obtained Institutional Review Board (IRB) approval, interpreted the data, and critically reviewed and revised the manuscript. V. A. M. S., D. B. H., and J. C. S. fabricated the CandyCollect devices and prepared devices for human subject research, including designing experimental protocols/engineering designs, and critically reviewed and revised the manuscript. A. M. M. and X. S. designed the sample processing protocol and drafted, critically reviewed, and revised the manuscript. E. B. conceptualized the research, advised on study design and interpretation, and critically reviewed and revised the manuscript. A. B. T., G. P. D., E. R. W., and S. T. conceptualized the research, designed the study, oversaw study execution, interpreted the data, and critically reviewed and revised the manuscript. All authors approved the final manuscript as submitted and agreed to be accountable for all aspects of the work.

## Notes

**Source of funding:** This work was supported by National Institutes of Health grants (R21AI166120, R35GM128648 (the latter specifically supported some of the in-lab developments and procedure developments)); the Washington Research Foundation; the Camille and Henry Dreyfus Foundation; and an Alfred P. Sloan Research Fellowship. The content is solely the responsibility of the authors and does not necessarily represent the official views of the National Institutes of Health or other funding sources.

### Competing Interest Statement

Ashleigh B. Theberge, Xiaojing Su, Erwin Berthier, and Sanitta Thongpang filed patent 63/152,103 (International Publication Number: WO 2022/178291 Al) through the University of Washington on the CandyCollect oral sampling device. Ashleigh B. Theberge reports filing multiple patents through the University of Washington and receiving a gift to support research outside the submitted work from Ionis Pharmaceuticals. Erwin Berthier is an inventor on multiple patents filed by Tasso, Inc., the University of Washington, and the University of Wisconsin. Sanitta Thongpang has ownership in Salus Discovery, LLC, and Tasso, Inc. Erwin Berthier has ownership in Salus Discovery, LLC, and Tasso, Inc. and is employed by Tasso, Inc. However, this research is not related to these companies. Sanitta Thongpang, Erwin Berthier, and Ashleigh B. Theberge have ownership in Seabright, LLC, which will advance new tools for diagnostics and clinical research, potentially including the CandyCollect device. The terms of this arrangement have been reviewed and approved by the University of Washington in accordance with its policies governing outside work and financial conflicts of interest in research. The other authors have no conflicts of interest to disclose.

### Clinical Trial

ClinicalTrials.gov Identifier: NCT05175196

### Funding Statement

All phases of this study were supported by National Institutes of Health grants (R21AI166120, R35GM128648 (the latter specifically supported some of the in-lab developments and procedure developments)), the Washington Research Foundation, the Camille and Henry Dreyfus Foundation, and an Alfred P. Sloan Research Fellowship. The content is solely the responsibility of the authors and does not necessarily represent the official views of the National Institutes of Health or other funding sources.

### Author Declarations

The Health Sciences IRB of the University of Wisconsin-Madison gave ethical approval for this work.

### Summary of Updates

- Update the current version of abstract (based on the journal we resubmitted) - Add two figures to the manuscript - Change one author's last name (she just has a new one)

