## Supplemental materials for "Capture of Group A Streptococcus by Open-Microfluidic CandyCollect Device in Pediatric Patients"

**Table S1.** Survey questions for user feedback (children)

**Table S2.** Survey questions for user feedback (caregivers)

**Table S3.** Free response question

**Table S4.** Study Participant Characteristics

**Figure S1.** Schematic of the CandyCollect device dimensions. All dimensions are in mm

**Figure S2.** Wong-Baker FACES® Pain Rating Scale

**Figure S3.** Pooled and individual cycle threshold (Ct) values of samples collected from mouth swabs and CandyCollect devices

**Figure S4.** Standard curves for the *S. pyogenes* qPCR assays, and qPCR amplification plots of standard curve and negative controls from a representative of three qPCR plates

**Table S1.** Survey questions for user feedback (children)

| | | | |
|---|---|---|---|
| 1 | Circle a number below to show how much pain (discomfort) you felt during the THROAT SWAB. [Children were shown the Wong-Baker FACES® Pain Rating Scale, refer to Figure S2.] | 0 | No hurt |
| | | 2 | Hurts Little Bit |
| | | 4 | Hurts Little More |
| | | 6 | Hurts Even More |
| | | 8 | Hurts Whole Lot |
| | | 10 | Hurts Worst |
| | | 99 | Declined to answer |
| 2 | Circle a number below to show how much pain (discomfort) you felt during the MOUTH SWAB. [Children were shown the Wong-Baker FACES® Pain Rating Scale, refer to Figure S2.] | 0 | 0 No hurt |
| | | 2 | 2 Hurts Little Bit |
| | | 4 | 4 Hurts Little More |
| | | 6 | 6 Hurts Even More |
| | | 8 | 8 Hurts Whole Lot |
| | | 10 | 10 Hurts Worst |
| | | 99 | Declined to answer |
| 3 | Circle a number below to show how much pain (discomfort) you felt during the CANDYCOLLECT (lollipop). [Children were shown the Wong-Baker FACES® Pain Rating Scale, refer to Figure S2.] | 0 | 0 No hurt |
| | | 2 | 2 Hurts Little Bit |
| | | 4 | 4 Hurts Little More |
| | | 6 | 6 Hurts Even More |
| | | 8 | 8 Hurts Whole Lot |
| | | 10 | 10 Hurts Worst |
| | | 99 | Declined to answer |
| 4 | If you needed to have another test for strep throat next week, which would you prefer? | 1 | Throat Swab |
| | | 2 | Mouth Swab |

| | | |
|---|---|---|
| | | 3 CandyCollect (lollipop)<br>99 Declined to answer |
| 5 | Would you be willing to do the CandyCollect (lollipop) at home? | 1 Yes<br>2 No<br>99 Declined to answer |
| 6 | Was the CandyCollect (lollipop) easy to suck on? | 1 1 Very Easy<br>2 2<br>3 3<br>4 4<br>5 5 Very Hard<br>99 Declined to answer |
| 7 | Did you like the taste of the CandyCollect (lollipop)? | 1 1 Really like it<br>2 2<br>3 3<br>4 4<br>5 5 Don't like it<br>99 Declined to answer |

**Table S2.** Survey questions for user feedback (caregivers)

<table>
<tr><td>1</td><td>Think back to the moment when your child was having the THROAT SWAB done by the clinic staff. Please provide your impressions of the THROAT SWAB by marking your impression on the scale between Pleasant and Unpleasant.</td><td><table><tr><td>1</td><td>1 - Pleasant</td></tr><tr><td>2</td><td>2</td></tr><tr><td>3</td><td>3</td></tr><tr><td>4</td><td>4</td></tr><tr><td>5</td><td>5</td></tr><tr><td>6</td><td>6</td></tr><tr><td>7</td><td>7 - Unpleasant</td></tr></table></td></tr>
<tr><td>2</td><td>How would you rate the THROAT SWAB overall?</td><td>Slider (number, Min: -5, Max: 5)<br>Slider labels: Bad, Good</td></tr>
<tr><td>3</td><td>Think back to the moment when your child was having the MOUTH SWAB done by the clinic staff. Please provide your impressions of the MOUTH SWAB by check marking your impression on the scale between Pleasant and Unpleasant.</td><td><table><tr><td>1</td><td>1 - Pleasant</td></tr><tr><td>2</td><td>2</td></tr><tr><td>3</td><td>3</td></tr><tr><td>4</td><td>4</td></tr><tr><td>5</td><td>5</td></tr><tr><td>6</td><td>6</td></tr><tr><td>7</td><td>7 - Unpleasant</td></tr></table></td></tr>
<tr><td>4</td><td>How would you rate the MOUTH SWAB overall?</td><td>Slider (number, Min: -5, Max: 5)<br>Slider labels: Bad, Good</td></tr>
<tr><td>5</td><td>Think back to the moment when your child was having the CandyCollect (lollipop) done by the clinic staff . Please provide your impressions of the CandyCollect (lollipop) by checkmarking your impression on the scale between Pleasant and Unpleasant.</td><td><table><tr><td>1</td><td>1 - Pleasant</td></tr><tr><td>2</td><td>2</td></tr><tr><td>3</td><td>3</td></tr><tr><td>4</td><td>4</td></tr><tr><td>5</td><td>5</td></tr><tr><td>6</td><td>6</td></tr></table></td></tr>
</table>

| | | |
|---|---|---|
| | | 7 \| 7 - Unpleasant |
| 6 | How would you rate the CandyCollect (lollipop) overall? | Slider (number, Min: -5, Max: 5)<br>Slider labels: Bad, Good |
| 7 | If your child needed to have another test for strep throat next week, which would you prefer? | 1 \| Throat Swab<br>2 \| Mouth Swab<br>3 \| CandyCollect (lollipop)<br>99 \| Declined to answer |
| 8 | What do you think is the most suitable sampling method for children in general? | 1 \| Throat Swab<br>2 \| Mouth Swab<br>3 \| CandyCollect (lollipop)<br>99 \| Declined to answer |
| 9 | Would you be willing to have your child do the CandyCollect (lollipop) at home? | 1 \| Yes<br>2 \| No<br>99 \| Declined to answer |
| 10 | Does the CandyCollect (lollipop) look appealing (the color, the overall appearance)? | 1 \| 1 Really like it<br>2 \| 2<br>3 \| 3<br>4 \| 4<br>5 \| 5 Don't like it<br>99 \| Declined to answer |
| 11 | Would you recommend these CandyCollect (lollipop) to children ages 5 and above? | 1 \| Yes<br>2 \| No<br>99 \| Declined to answer |

**Table S3.** Free response question

| Please explain your response | How can we improve our CandyCollect (lollipop) for children? Provide any suggestions |
|---|---|
| Not scary, pleasant color, tasty, soothing on throat | Shorter time, other flavors, not sure about the sugar free sweetener |
| Child friendly, appealing to children. | No ideas |
| It takes longer than a swab but it is way more pleasant and fun for the child. | I think it's a great idea. Only complaint is that it takes a little while-but my child didn't mind! |
| Compared to throat swab, more appealing/less scary. Could help kids be less afraid to go to the doctor in general. | Consider making handle white instead of clear to make it look more like a lollipop. |
| My child is 10 yr old: he was in tears on the way here dreading the throat swab test. The candy collect would not only provide test results but also perhaps soothe his sore throat. | My only suggestion would be having a few flavors? |
| It seems fun for kids. | N/A |
| N/A | No suggestions |
| Kids like lollipops so not scary | Not take as long |
| They would be old enough to follow instructions and not have to worry of choking. They would also enjoy it. | N/A |
| Great, non-invasive test for an already painful ailment. | If it goes forward-esp over the counter-very easy, detailed instructions are helpful. |
| N/A | N/A |
| The CandyCollect was easy and fast; he seemed to enjoy it and there was no pain-big plus!!! :) | Different flavors for kids to choose from |
| My child seemed to find it far more pleasant than the throat swab. | I can't think of any suggestions. |
| N/A | Avoiding food dye would be beneficial for many children. |
| This is a simple, appealing test for kids: the color, size, and flavor all seem like a lollipop. | Make sure that parents know whether it includes red artificial food dye. Some parents avoid red dye #40 and similar additions. Parents might feel reassured knowing the ingredients and whether vegetable dye is used. |
| Seems like the candy would be way more appealing than having a throat swab. | Variety of flavors |

| | |
|---|---|
| Great alternative to throat swab | Quicker collection |
| Easy for kids to suck on as it tastes good and kids like suckers | None |
| Child was excited to do the test. | Nothing |
| No discomfort, especially compared to throat swab. | None |
| I feel based on the time for it dissolve that it would be better for kids 5 and older. I think that is a great age to be able to do that and wait. My son has a strong gag reflex so I think this was great. | I don't have any suggestions for improvement. I think it was a great way to collect a sample and I really hope strep can be detected this way. |
| My child liked the flavor, enjoyed the concept. It takes a bit longer than the mouth swab though. | Maybe have it dissolve in less time. |
| Easy, non-invasive | Quicker process |
| The mouth swab was easy, but if it's not effective the Candycollect was just as good! | Maybe shorter suck time?! |
| It is more pleasant than throat swab by far-but was time consuming. | No suggestions |
| Much easier than throat swab. | N/A |
| Really liked the concept of the CandyCollect but the time was a little long. The mouth swab is quick and easy even though no flavor. | N/A |
| Great way to collect in a fun way for kids. | My only concern would be the length of time and how much swallowing they have to do if their throat is very sore, and if they aren't feeling well. |
| N/A | I don't have any feedback. |
| If it didn't take as long | Offer a variety of flavors (Grape, Bubblegum), too long to do test |

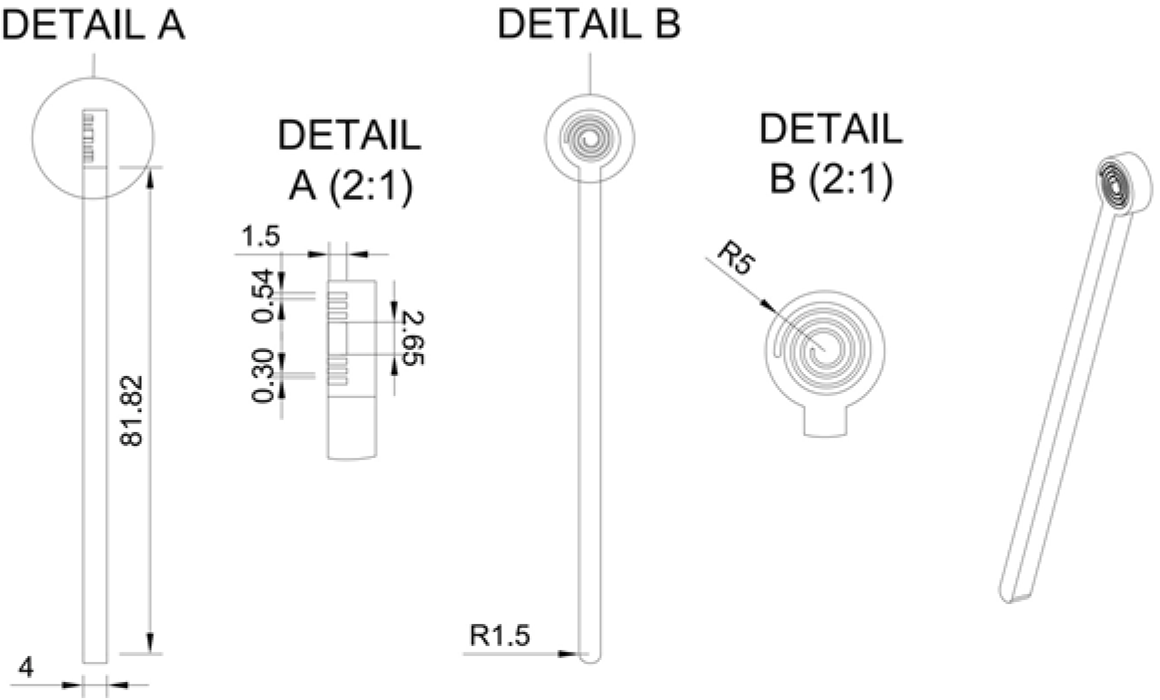


**Figure S1.** Schematic of the CandyCollect device dimensions. All dimensions are in mm. This figure is reproduced from Tu et al. [1] (Figure S1) with permission from the ACS publication (*Analytical Chemistry*).

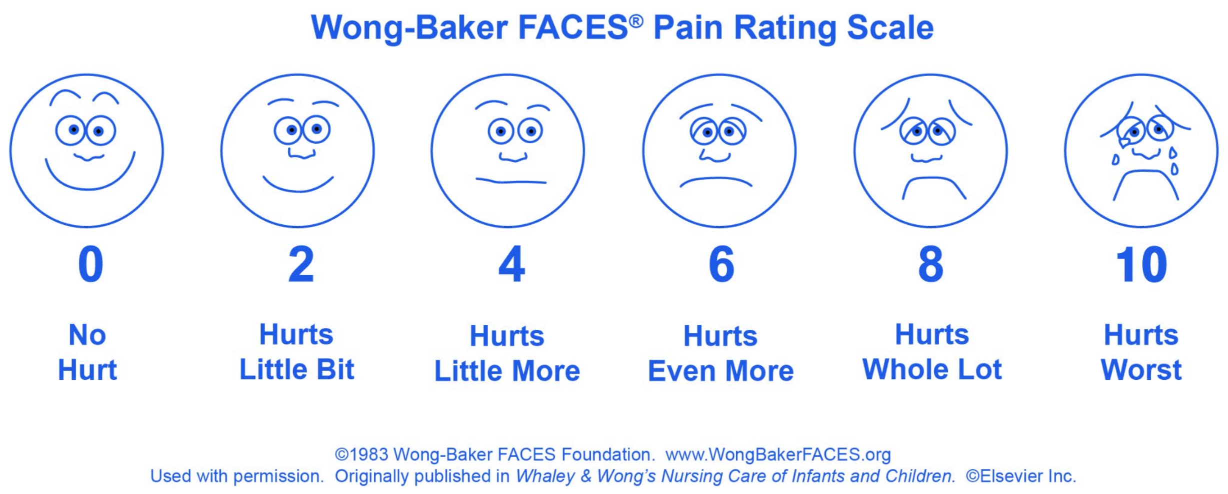


**Figure S2.** Wong-Baker FACES® Pain Rating Scale.[2] Face 0 represents does not hurt at all; face 2 represents hurts just a little bit; face 4 represents hurts a little bit more; face 6 represents hurts even more; face 8 represents hurts a whole lot; face 10 represents hurts as much as you can imagine, although you do not have to be crying to have this worst pain.

**Table S4.** Study Participant Characteristics (Child Participants)

| Characteristic | In this study No. (percentage) | In Dane County, Wisconsin[3] |
|---|---|---|
| Mean age | 8.8 ± 2.5 years | |
| Female, n (%) | 16 of 30 (53%) | 49.9% |
| Race, n (%) | | |
| American Indian or Alaska Native | 0 (0%) | 0.5% |
| Asian | 1 (3%) | 6.6% |
| Black or African American | 1 (3%) | 5.8% |
| Native Hawaiian or Other Pacific Islander | 0 (0%) | 0.1% |
| White | 28 (93%) | 84.1% |
| Ethnicity, n (%) | | |
| Hispanic or Latino | 1 (3%) | 7.0% |
| Not Hispanic or Latino | 29 (97%) | 78.0% |

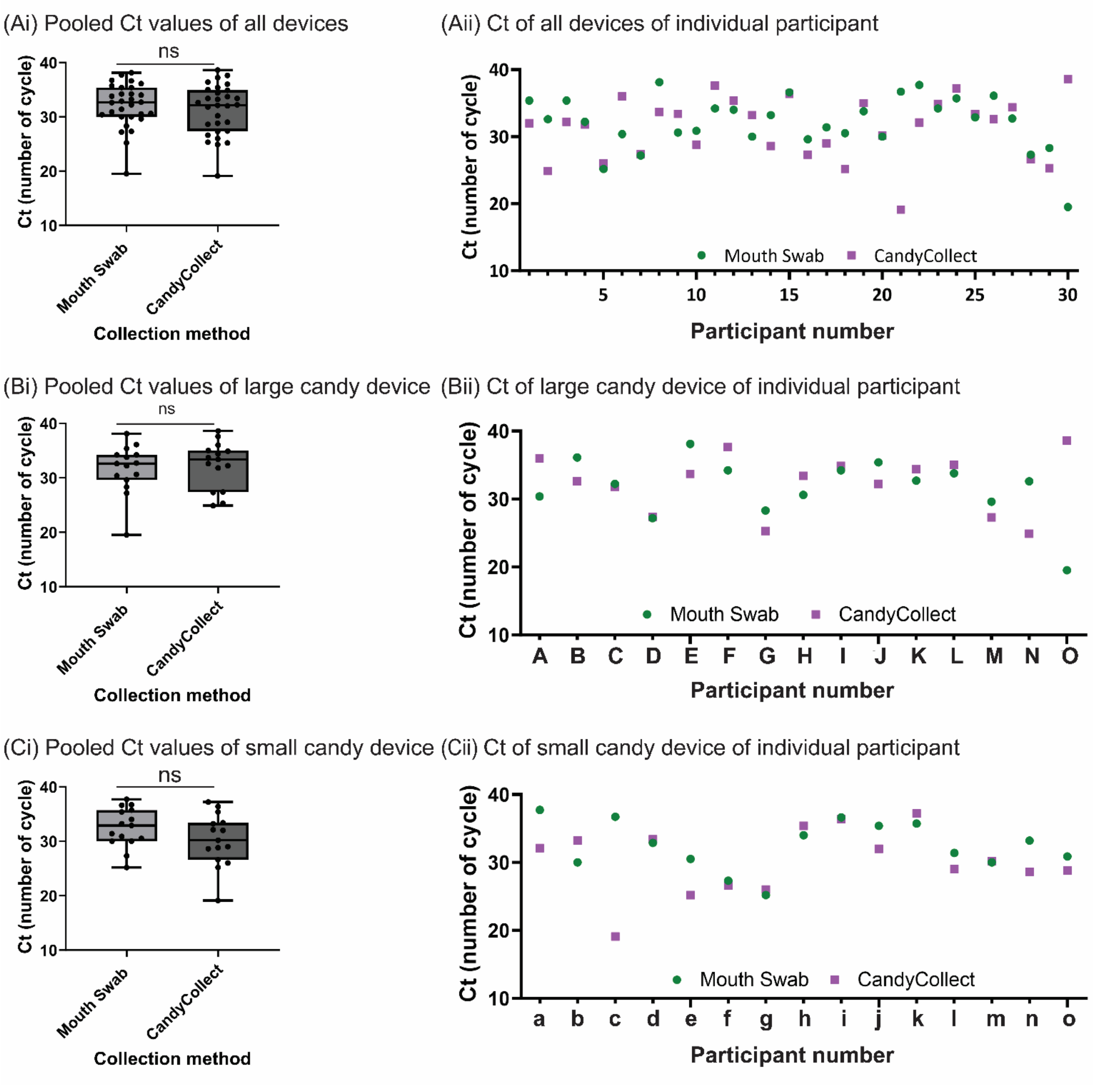


**Figure S3.** Pooled and individual cycle threshold (Ct) values of samples collected from mouth swabs and CandyCollect devices (A) all 30 child participants (B) from15 participants using the large CandyCollcet devices (Candy = 1.3g, green) and (C) from 15 participants using small CandyCollect devices (Candy = 0.5g, purple). Ct values from the two sampling methods are not significantly different (p>0.05). The participant numbers were randomly re-numbered.

**Method (This section is reproduced from Lee et al. [3] and Tu et al. [1])**

**Bacteria culture**

*Liquid media preparation*

For the THY liquid media, 30 g of Todd-Hewitt Broth (BD Bacto™ TH broth, Fisher Scientific, Cat# DF0492-17-6) and 2 g of Yeast Extract (United States Biological Corporation, Fisher Scientific, Cat# NC9796728) (THY) were added to 0.8 L distilled water and dissolved to completion. Additional distilled water was added for a total volume of 1 L. THY liquid media was autoclaved at 121 °C for 30 min, cooled to room temperature, and stored at 4 °C.

*Agar plate preparation*

7.5g agar (BD Difco™ Dehydrated Culture Media: Potato Dextrose Agar, Fisher Scientific, Cat# DF0013-17-6) was added to the 500 mL of liquid media, then autoclaved at 121 °C for 30 min. 15 mL of liquid media with agar was added to petri dishes, left to cool overnight, and stored at 4°C until needed for.

*S. pyogenes maintenance in agar plate*

Freeze-dried *S. pyogenes* was rehydrated with 1 mL liquid media, and then transferred to another conical tube containing 4.4 mL of liquid media. To maintain the bacteria, *S. pyogenes* were streaked on agar plates by sterile disposable inoculating loops (Globe Scientific, Fisher Scientific, Cat# 22-170-201). The agar plates were incubated at 37 °C with 5% carbon dioxide overnight, then stored at room temperature for up to 7 days.

**In-lab capture of bacteria on CandyCollect devices and mouth swab**

*Incubation of S. pyogenes in liquid media*

To ensure a pure culture, fresh *S. pyogenes* from agar plates were inoculated in liquid media and cultured at 37 °C with 5% carbon dioxide in the incubator one day prior to an experiment.

*Preparation of positive and negative controls*

After culturing overnight, the bacteria suspensions were homogenized with vortexing at a concentration of $1x10^4$ CFU/mL and added to each CandyCollect device at a volume of 50 µL and incubated for 10 minutes; for the mouth swab, the swab was soaked in homogenized bacteria suspensions for 10 seconds (PBS was used for device negative controls).

**Specimen Processing and Laboratory Analysis**

Research nurse stored the mouth swab (ESwab™) and CandyCollect samples at -20 °C for a few days after sampling at Madison, Wisconsin, and the samples were shipped to University of Washington using United Parcel Service (UPS) Next Day Air. Samples were stored at -80 °C before processing. All laboratory procedures were performed in accordance with Biosafety Level-2 laboratory practices and the University of Washington Site-Specific Bloodborne Pathogen Exposure Control Plan. To avoid unnecessary freeze-thaw cycles, mouth swab (ESwab™) samples were aliquoted into 20 µL aliquots and stored at -80 °C.

*Elution of S. pyogenes from CandyCollect devices*

The buffer used to elute bacteria captured on CandyCollect devices was phosphate buffered saline (PBS) (Gibco™, Cat# 10010023) with 5% Proteinase K (Thermo Scientific™, Cat# EO0491). 300 µL elution buffer and 100 µL of 0.1 mm Zirconia/Silica beads (BioSpec Products, Cat# 11079101Z) were added in 14 mL round bottom tubes (Corning, Falcon®, 352001) containing CandyCollect devices. After incubating the tubes at 37 °C for 10 min and vortexing for 50 s, CandyCollect devices were left in the elution buffer at 4 °C for 90 min. The bacteria suspension and beads were then transferred from the 14 mL round bottom tubes to 2 mL screw cap microtubes (ThermoFisher, Cat# 3490). The samples were beat-beaten in a MiniBeadBeater (BioSpec Products, Bartlesville, OK USA), and stored at -20 °C before analysis.

*DNA isolation from mouth swab samples*

DNA from mouth swab was isolated using MagMAX™ Total Nucleic Acid Isolation Kit (ThermoFisher Scientific, Cat# AM1840) according to the protocol "Disruption of liquid samples" supplied by the manufacturer. Briefly, 175 µL of aliquoted samples were transferred to each bead beating tube provided in the kit followed by the addition of 230 µL of Lysis/Binding solution. Bead beating was carried out via MiniBeadBeater (mentioned above) twice for 30 s, then each tube was centrifuged at 16,000 g for 3 min. Afterward, genomic DNA of *S. pyogenes* was isolated and enriched following the protocol stated above and quantified using qPCR with a detection limit of 5 fg.

*Isolation, purification, and enrichment of genomic DNA from S. pyogenes*

DNA was isolated from bacterial lysates using the MagMAX™ Total Nucleic Acid Isolation Kit (ThermoFisher Scientific, Cat# AM1840) according to the "Purify the nucleic acid" protocol supplied by the manufacturer. In brief, 115 µL of sample was added to the provided processing plate. 60 µL of 100% IPA was added to each well containing a sample and the plate was shaken for 1 min. 20 µL of bead mix was then added to each well, and the plate was shaken for 5 min to allow DNA to bind to the beads. Beads were captured using a magnetic 96-well separator (Thermofisher, Cat# A14179) and supernatant was discarded. Four washes (two using Wash Solution 1 and additional two using Wash Solution 2 provided by the kit) were performed with shaking for 1 min each and supernatant was discarded between each wash. After final wash, beads were dried and then 23 µL of 65°C elution buffer was added to each sample to elute DNA from the beads. By using these methods, DNA was five-fold concentrated compared to the unprocessed bacterial lysates. The purified bacterial genomic DNA was used as a template in the qPCR assay.

*Quantitative PCR assay for detection S. pyogenes*

The details for the qPCR assay for *S. pyogenes* followed the protocol from our previous paper.[1] Briefly, the primers/probe sequences for *spy1258* qPCR detection of *S. pyogenes* in our assay were: the forward primer: 5'-GCA CTC GCT ACT ATT TCT TAC CTC AA-3'; the reverse primer: 5'-GTC ACA ATG TCT TGG AAA CCA GTA AT-3'; the probe sequence: 5'-FAM-CCG CAA C"T"C ATC AAG GAT TTC TGT TAC CA-3'-SpC6, "T" = BHQ1.1 The primers were ordered from IDT, the probe was ordered from MilliporeSigma. The 25 µL reaction volume included 10 µL of DNA template and 15 µL PerfeCTa® qPCR ToughMix with primers/probe in the qPCR assay. The final concentrations of both forward and reverse primers were 300 nM; the probe concentration was 100 nM. To quantify the DNA concentrations of samples, 1:10 serial dilution of purified genomic DNA ranging from 5 ng to 5 fg were used as standards for each plate. No-template controls (NTC) for qPCR and device negative controls were also added to the plates. Amplification and detection were performed in 96-well PCR plates using CFX connect Real-Time PCR Detection System (Bio-Rad Laboratories, Hercu-les, CA, USA) in technical duplicate using the following protocol: 95 °C for 5 min followed by 40 cycles of 15 s at 95 °C and 30 s at 60 °C. The samples were considered positive when the Ct value is within the Ct of the standard curve.

*Threshold cycle (Ct) evaluation for positive results of human subject samples*

To evaluate qPCR efficiency and specificity, a 1:10 serial dilution of purified genomic DNA (5 ng -5 fg) isolated from *S. pyogenes* was used to construct a standard curve (Figure S6A and S6B) along with the following three different negative controls: 1. Device control - experiments were performed with CandyCollect devices running through all the procedures using PBS, instead of *S. pyogenes* suspension. 2. Other bacterial species controls - experiments were performed with *Streptococcus mutans* and *Staphylococcus aureus*. 3. qPCR control - elution buffer from the MagMAX™ Total Nucleic Acid Isolation Kit was used as non-template control (Figure S6C). While all DNA concentrations of DNA standard generate normal PCR amplification curves, all negative controls either did not show any amplification curve or resulted in the Ct value higher than 40, indicating PCR specificity. Furthermore, the efficiency of the qPCR standard is within 100-103 %. After resolving on 3% agar gel, a single band was shown for qPCR products from all tested concentrations of standards, further validating the specificity of qPCR amplification (Lee et al. Figure S4).[3] Clinical samples were run across three PCR plates, and a standard curve was run separately on each plate. The Ct values for the lowest concentration of DNA standards (5 fg) were 37.96, 38.88, and 38.23, and we compared the Ct values of the samples with the lowest Ct value of standard curve on the same plate. This number was used as a cut-off number for differentiating positive and negative detection. Any Ct value below or equal to Ct values for the lowest concentration of DNA standards (5 fg) on the same plate was considered positive, and all clinical samples were below the cycle threshold and therefore positive.

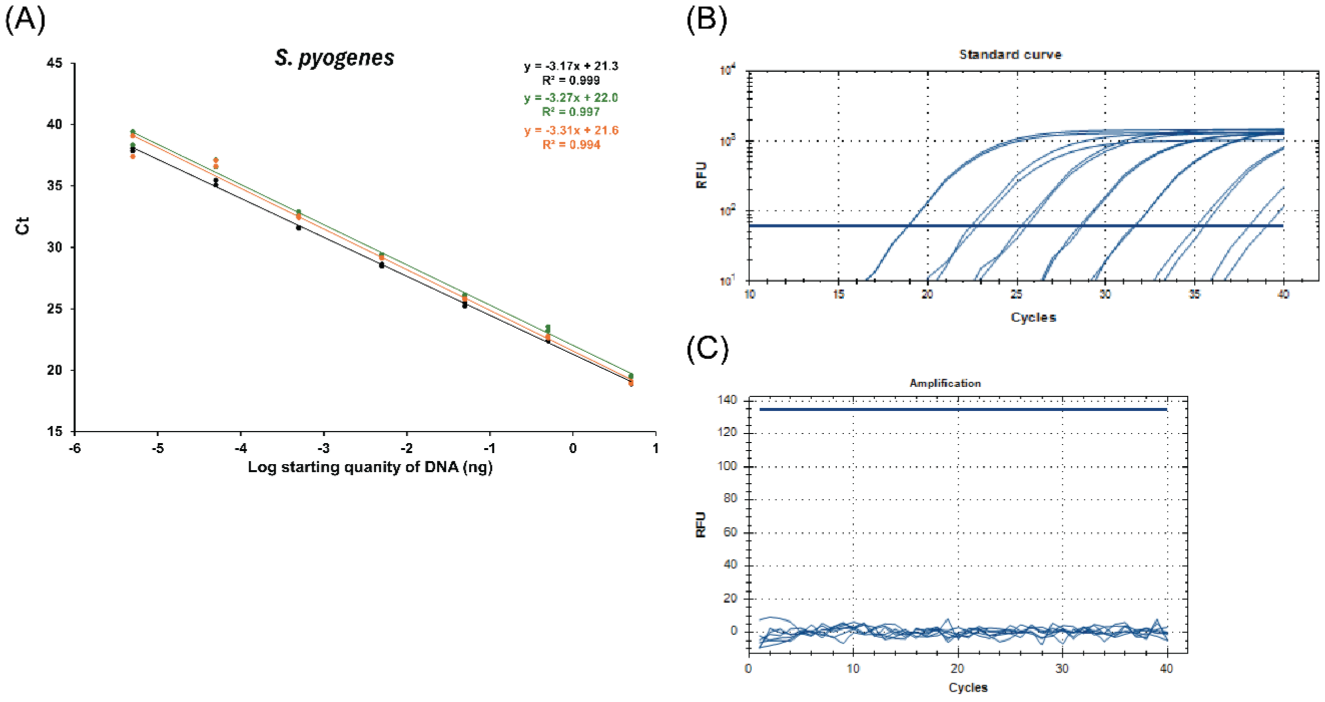


**Figure S4.** (A) Standard curves for the *S. pyogenes* qPCR assays. 1:10 serial dilutions of genomic DNA ranging from 5 ng to 5 fg were used as templates for qPCR. Each dot represents one technical duplicate (in cases where one point is visible the duplicates were identical). (B) The qPCR amplification plots of standard curve and (C) the qPCR amplification plots of negative controls from a representative of three qPCR plates.
